# COVID-19 vaccines that reduce symptoms but do not block infection need higher coverage and faster rollout to achieve population impact

**DOI:** 10.1101/2020.12.13.20248142

**Authors:** David A. Swan, Chloe Bracis, Holly Janes, Mia Moore, Laura Matrajt, Daniel B. Reeves, Eileen Burns, Deborah Donnell, Myron S. Cohen, Joshua T. Schiffer, Dobromir Dimitrov

**Author notes:** **Corresponding author:** Dobromir Dimitrov, Vaccine and Infectious Disease Division, Fred Hutchinson Cancer Research Center, 1100 Fairview Ave N., M2-C200, P.O. Box 19024, Seattle, WA 98109-1024, USA. These authors contributed equally to the work (David A. Swan, Chloe Bracis). These authors contributed equally to the work (Joshua T. Schiffer, Dobromir Dimitrov).

## Abstract

**Background:** Several COVID-19 vaccine candidates are in the final stage of testing. Interim trial results for two vaccines suggest at least 90% efficacy against symptomatic disease (VE_DIS_). It remains unknown whether this efficacy is mediated predominately by lowering SARS-CoV-2 infection susceptibility (VE_SUSC_) or development of symptoms after infection (VE_SYMP_). A vaccine with high VE_SYMP_ but low VE_SUSC_ has uncertain population impact.

**Methods:** We developed a mathematical model of SARS-CoV-2 transmission, calibrated to demographic, physical distancing and epidemic data from King County, Washington. Different rollout scenarios starting December 2020 were simulated assuming different combinations of VE_SUSC_ and VE_SYMP_ resulting in up to 100% VE_DIS_ with constant vaccine effects over 1 year. We assumed no further increase in physical distancing despite expanding case numbers and no reduction of infectivity upon infection conditional on presence of symptoms. Proportions of cumulative infections, hospitalizations and deaths prevented over 1 year from vaccination start are reported.

**Results:** Rollouts of 1M vaccinations (5,000 daily) using vaccines with 50% VE_DIS_ are projected to prevent 30%-58% of infections and 38%-58% of deaths over one year. In comparison, vaccines with 90% VE_DIS_ are projected to prevent 47%-78% of the infections and 58%-77% of deaths over one year. In both cases, there is a greater reduction if VE_DIS_ is mediated mostly by VE_SUSC_. The use of a “symptom reducing” vaccine will require twice as many people vaccinated than a “susceptibility reducing” vaccine with the same 90% VE_DIS_ to prevent 50% of the infections and death over one year. Delaying the start of the vaccination by 3 months decreases the expected population impact by approximately 40%.

**Conclusions:** Vaccines which prevent COVID-19 disease but not SARS-CoV-2 infection, and thereby shift symptomatic infections to asymptomatic infections, will prevent fewer infections and require larger and faster vaccination rollouts to have population impact, compared to vaccines that reduce susceptibility to infection. If uncontrolled transmission across the U.S. continues, then expected vaccination in Spring 2021 will provide only limited benefit.

## Introduction

SARS-CoV-2 has infected more than 69 million people around the world as of December 10, 2020, causing close to 1.6 million deaths.^1^ With the onset of winter in the Northern Hemisphere, cases are surging particularly in Europe and North America. The need for a vaccine is the most urgent public health priority in recent human history.^2,3^

Two mRNA-based vaccines have already demonstrated efficacy greater than 90% and several more vaccine candidates are in the final stage of testing.^4,5^ The primary objective in each of these studies is to evaluate vaccine efficacy against virologically-confirmed symptomatic SARS-CoV-2 infection, i.e. COVID-19 disease, denoted VE_DIS_. The COVID-19 endpoint is defined by protocol-specified criteria for signs and symptoms consistent with COVID-19 and a confirmatory SARS-CoV-2 PCR test. Asymptomatic cases are therefore not captured in the primary endpoint of these trials, though seroconversion data will ultimately be available for all participants and potentially used to assess the proportion of asymptomatic infections in the active and placebo arms.

The population impact of a vaccine is not fully captured by its efficacy against disease; several other types of vaccine effects may have as much or greater influence.^6^ First, the extent to which a COVID-19 vaccine reduces the likelihood of acquiring SARS-CoV-2 upon exposure, i.e.. vaccine efficacy on susceptibility (VE_SUSC_), will have a dramatic effect on population impact. A vaccine’s effect on reducing susceptibility may be full abrogation of infection (an ‘all or none’ mechanism) or partial, whereby the per-exposure susceptibility to infection is reduced (a ‘leaky’ mechanism).^7,8^ Alternatively, or in combination, a vaccine might reduce the likelihood of symptoms upon infection (VE_SYMP_) and lead to subclinical (asymptomatic) infection with viral shedding that might still allow ongoing transmission. Critically, VE_SYMP_ would also contribute to observed VE_DIS_. Finally, a vaccine may decrease the infectiousness of individuals who become infected (VE_INF_). Fully appreciating a vaccine’s population impact requires estimating all of these vaccine effects. The task of evaluating the overall effectiveness of a vaccine is further complicated if the magnitude and durability of some or all of the vaccine effects varies with age, force of infection, or other host or virologic factors.^9-11^

A specific concern for COVID-19 vaccines is that a vaccine with high efficacy against COVID-19 disease (VE_DIS_) but low efficacy against SARS-CoV-2 infection (VE_SUSC_), would predominantly convert symptomatic infections to asymptomatic infections, but if the vaccine does not reduce infectiousness (VE_INF_ =0), it could in theory lead to increased spread of SARS-CoV-2.^12,13^ This is especially relevant in settings such as the US where the testing and diagnosis of infection is primarily symptom-driven; individuals with asymptomatic infection are less likely to be diagnosed and therefore may spread infection more readily than diagnosed and socially-isolated symptomatic individuals. The concern is heightened by considerable evidence pointing to the role that asymptomatic and pre-symptomatic cases play in transmission of SARS-CoV-2, with peak infectiousness typically occurring prior to the onset of symptoms.^14-16^ A vaccine that shifted the burden of infection to largely asymptomatic infections, without reducing their infectiousness, could theoretically worsen the pandemic. The Pfizer and Moderna vaccines are reported to have ∼95% VE_DIS_ with VE_SUSC_ and VE_SYMP_ remaining unknown. Unfortunately, VE_INF_ for these vaccines is not estimable from the completed trials.

Mathematical models are useful tools to simulate epidemic dynamics and several models have already projected the population impact of hypothetical COVID-19 vaccines.^17-26^ While some of them have included vaccines with different vaccine effects, this work has mostly revolved around vaccine prioritization groups.^17,18^ Gallagher et al^22^ investigated the population impact of a vaccine with low VE_DIS_ but high VE_INF_ but none have specifically assessed the population impact of “symptom averting” vaccines with high VE_DIS_ but low VE_SUSC_ and VE_INF_. As a result, the modeling projections of achieving more than 80% population effectiveness with 40% coverage may be overly optimistic.^24^ Moreover, most of the previous work has not considered vaccination rollouts initiated in the middle of epidemic wave which may reduce the overall benefits from the vaccination.

Employing a mathematical model calibrated to King County, WA, this study addresses the specific question if a vaccine which reduces the risk of symptomatic disease but has low efficacy in reducing susceptibility to infection and little to no impact on infectiousness may adversely impact the epidemic. More generally, we analyze the expected population benefits of a COVID-19 vaccine with demonstrated efficacy in reducing COVID-19 disease under varying combinations of vaccine effects (VE_SUSC_ and VE_SYMP_) and under different scenarios in terms of timing and population coverage of vaccine rollout. Our results suggest that the concern for increased transmission due to rollout of COVID-19 vaccines that have at least 50% VE_DIS_ and convert symptomatic to asymptomatic infections (low VE_SUSC_ and high VE_SYMP_) is not supported by the evidence, but that vaccines that do reduce the risk of infection (high VE_SUSC_ and low VE_SYMP_) are expected to have substantially greater population impact. We also demonstrate the importance of rolling out the vaccine fast and early, before an epidemic outbreak which is highly relevant to the current epidemic situation given the spike in the confirmed cases locally and nationwide.

## Methods

### Model description

We modified a previously developed deterministic compartment model which describes the epidemic dynamics in King County, WA^27^ to allow for vaccine rollouts. Our model (**Fig 1** and **Fig S1**) stratifies the population by age (0-19 years, 20-49 years, 50-69 years, and 70+ years), infection status (susceptible, exposed, asymptomatic, pre-symptomatic, symptomatic, recovered), clinical status (undiagnosed, diagnosed, hospitalized) and vaccination status.

**Figure 1.**
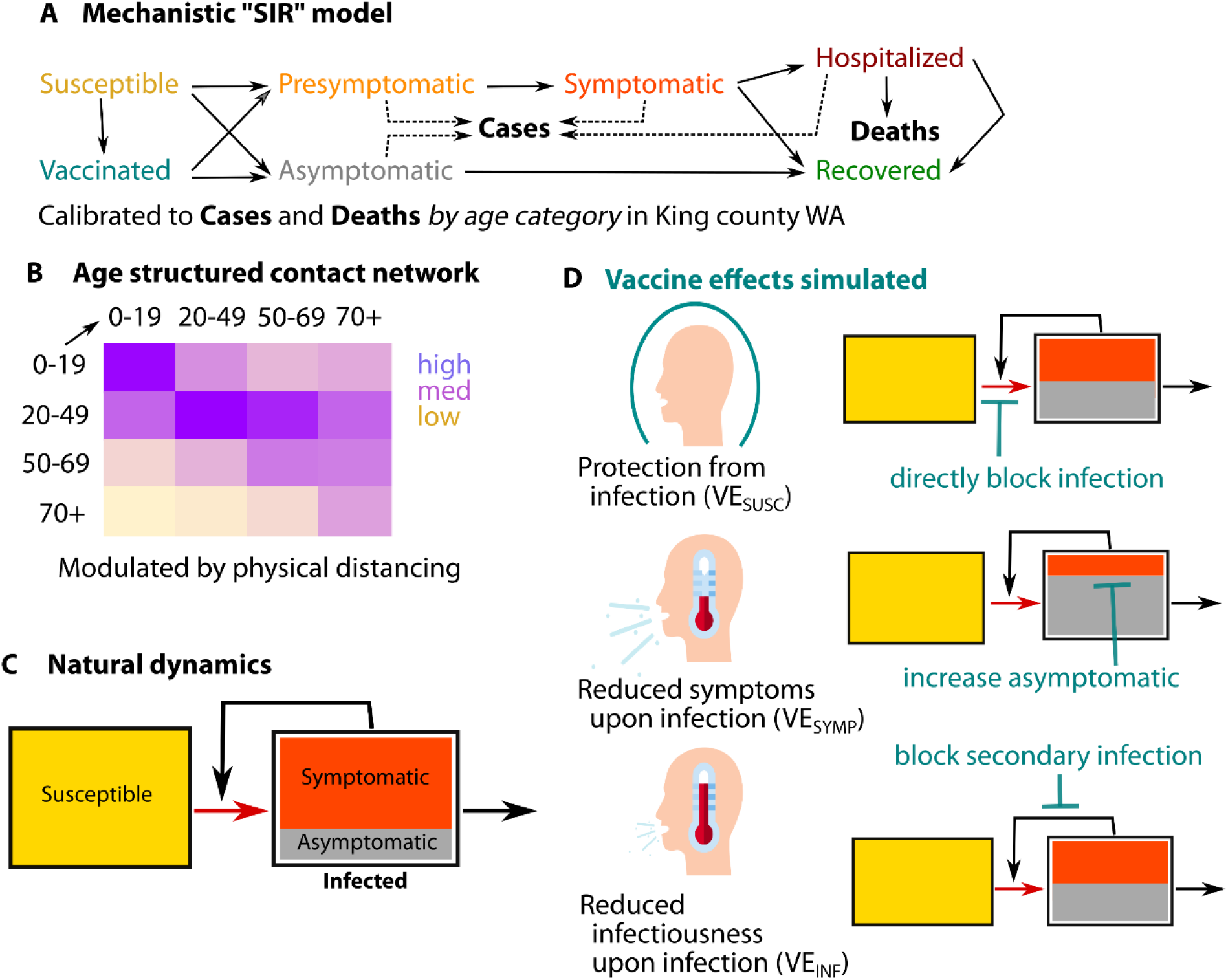
Schematic of the modeling analysis. A) Diagram of the population transmission model with solid lines representing the flows of individuals between compartments and dotted lines indicating compartments which contribute to diagnosed cases; B) Mixing matrix between age groups showing proportions of contacts each group (row) has with the other groups (columns); C) Simplified diagram of the SARS-CoV2 transmission (red arrow) and the resulting force of infection (black arrow) in absence of vaccine and D) the potential vaccine effects on the transmission and the force of infection

In our main scenario we assume that 20% of infections are asymptomatic reflecting the estimated proportion of SARS-CoV2 infections without symptoms in a published meta-analysis based on 79 studies.^15^ We also assume that asymptomatic people are as infectious as symptomatic individuals but missing the highly infectious pre-symptomatic phase. As a result, the relative infectiousness of individuals who never express symptoms is 56% of the overall infectiousness of individuals who develop symptomatic COVID-19 infection. This estimate falls between the 35% relative infectiousness estimated in the meta-analysis^15^ and the current best estimate of 75% suggested by the CDC in their COVID-19 pandemic planning scenarios^28^. We explore alternative scenarios in which the difference in overall infectiousness between asymptomatic and symptomatic cases is smaller (asymptomatic infections are only 28% less infectious than symptomatic cases) to assess the importance of this assumption to the presented results.

The forces of infection, representing the risk of the susceptible individuals to acquire infection (transition from susceptible to exposed), are differentiated by age of the susceptible individual, the contact matrix (proportion of contacts with each age group), infection and treatment status (asymptomatic, pre-symptomatic, symptomatic, diagnosed and hospitalized cases) of the infected contacts, and the time-dependent reduction of transmission due to physical distancing measures (work from home, closing non-essential businesses, banning large gathering, etc.) applied in the area during 2020.

The model is parameterized with local demographic and contact data from King County, WA and calibrated to local case and mortality data using transmission parameter ranges informed from published sources **(Figs S2** and **S3)**. ^29-32^ We used a genetic algorithm (NSGA-II multivariate optimization algorithm in the mco R package) and Monte Carlo filtering to select 100 calibrated parameter sets on epidemic start, transmission and hospitalization rates by age and physical isolation as a function of symptoms and diagnosis which reproduce the data within pre-specified tolerances. We validated our assumptions on the diagnostic rates of symptomatic and asymptomatic cases during “reopening” period against independent data not used for calibration (cases and deaths after the calibration period), comparison to expert predictions, and independent region-specific modeling projections^33,34^. As a result, we obtained an ensemble of plausible epidemic trajectories with variable projections of the timing and magnitude of the winter epidemic peak suitable to explore the uncertainty in the vaccine effectiveness predictions associated with background epidemic conditions. We also explored additional scenarios in which only symptomatic cases get tested and diagnosed to assess the importance of this assumption for the projected impact of symptom averting vaccines. A full description of the model can be found in the Supplement.

### Vaccination scenarios

We consider several vaccine efficacy profiles (**Table 1**) with respect to vaccine effects on susceptibility (VE_SUSC_), defined as reduction of the probability of acquiring infection upon exposure, and vaccine effects on symptomatology (VE_SYMP_), defined as the reduction in the probability to develop symptoms upon infection. In combination, these two parameters determine the expected reduction in the likelihood to develop symptomatic diseases upon exposure (VE_DIS_) as described in **Table 1**. Namely, we simulate and compare two profiles (Vaccine 1 and Vaccine 2) satisfying the WHO and FDA requirement for COVID-19 vaccines to be at least 50% efficacious reducing the risk of symptomatic disease (50% VE_DIS_)^35,36^, and two profiles (Vaccine 3 and Vaccine 4) with efficacy against COVID-19 disease (90% VE_DIS_) comparable to the results reported on the Pfizer and Moderna vaccines.^5^ The values explored for VE_SUSC_ and VE_SYMP_ allow us to clearly differentiate “susceptibility reducing” vaccines which actively reduce the risk to acquire infection (Vaccine 2 and 4) from “symptom reducing” vaccines which mostly convert symptomatic infections to asymptomatic (Vaccine 1 and 3). Since our goal is to investigate if a vaccine, meeting success criteria for efficacy against disease but with unclear efficacy against infection could theoretically increase transmission, we assume no vaccine effects on the infectiousness (VE_INF_=0) for either asymptomatic or symptomatic infections. However, all of the simulated vaccine profiles shift some symptomatic infections to asymptomatic infections, which are less infectious under the assumptions of the model. As a result, some degree of reduction of infectiousness due to vaccination occurs in all simulated scenarios.

**Table 1.** Vaccine efficacy profiles used in the main analysis. All vaccine profiles correspond to 50% reduction in the likelihood to develop symptomatic disease upon exposure (VE_DIS_).

| Vaccine efficacy profiles | Vaccine 1 | Vaccine 2 | Vaccine 3 | Vaccine 4 |
| --- | --- | --- | --- | --- |
| Reduction of susceptibility upon exposure ($VE_{SUSC}$) | 0.1 | 0.4 | 0.1 | 0.8 |
| Reduction of the risk of symptomatic disease upon acquisition ($VE_{SYMP}$) | 0.444 | 0.167 | 0.889 | 0.5 |
| Reduction of infectiousness upon acquisition ($VE_{INF}$) | 0 | 0 | 0 | 0 |
| Reduction of the risk of symptomatic disease upon exposure ($VE_{DIS}$)=$1-(1-VE_{SUSC})(1-VE_{SYMP})$ | 0.5 | 0.5 | 0.9 | 0.9 |

In our main scenario we assume 5000 vaccinations a day starting Dec 1, 2020 and continuing for 200 days (∼7 months) until 1,000,000 individuals are vaccinated, enough to cover ∼45% of the population. Our vaccination rate and population coverage are less optimistic compared to the recent prognosis by the Head of Operation Warp Speed who suggested that 70% of the U.S. population could be vaccinated by May.^37^ Alternative scenarios with delayed vaccination start dates and 200,000-2,000,000 total vaccinated (10%-90% population coverage) are also explored. Vaccinations are proportionally distributed across age groups without excluding individuals with prior SARS-CoV-2 infection. Vaccinated individuals are assumed to continue with the same level of physical interactions as unvaccinated, i.e. there is no behavioral disinhibition. We don’t explicitly model two-dose vaccine regimens but assume immediate immunization at the time of vaccination.

### Metrics of interest

Population effectiveness is estimated as: i) proportion of cumulative infections prevented; ii) proportion of cumulative hospitalizations prevented iii) proportion of cumulative deaths prevented. Reductions in maximum daily infections, hospitalizations and deaths over 1 year from the vaccination start date are also evaluated. The uncertainty in the estimates is presented as 80% uncertainty interval (UI) based on simulations with the 100 calibrated parameter sets per scenario.

## Results

### Base-case epidemic conditions

In the absence of a vaccine and further lockdown efforts over the next year to stem a surge in cases, the set of acceptable simulations selected in the calibration process result in epidemic outbreaks reaching a maximum of 5900 (80%UI: 3700-8300) new infections, 390 (80%UI: 230-550) new hospitalizations and 99 (80%UI: 65-138) deaths daily at their transmission peak expected between January and July 2021. As of Dec. 1, 2020, the model projects that 4.2% (80%UI: 1.4%-8.5%) of the population in King County will be either infected or recovered while 785,000 (80%UI: 647,000-914,000) cumulative infections and 13,500 (80%UI: 10900-16200) cumulative deaths are predicted by Dec 2021 if no vaccine becomes available. Most new infections and deaths will have accrued before June 2021 with saturation due to natural herd immunity and ongoing non-pharmaceutical measures.

### Population impact of vaccines reducing the risk of COVID-19 disease by 50%

A “symptom reducing” vaccine with 50% VE_DIS_ with no effects on the infectiousness of breakthrough infections (Vaccine 1: moderate VE_SYMP_, low VE_SUSC_) is projected to prevent 33% of the infections and 40% of the deaths over a 1-year period. The model projects that the use of Vaccine 1 will reduce the cumulative number of infections by 320,000 to approximately 560,000 and the cumulative number of deaths by almost 4,000 to approximately 8600 (**Fig.2**, Vaccine 1). Vaccine 1 is projected to reduce the maximum number of daily infections and burden on the health system in terms of expected hospitalization by approximately 35% **(Fig S4)**.

**Figure 2.**
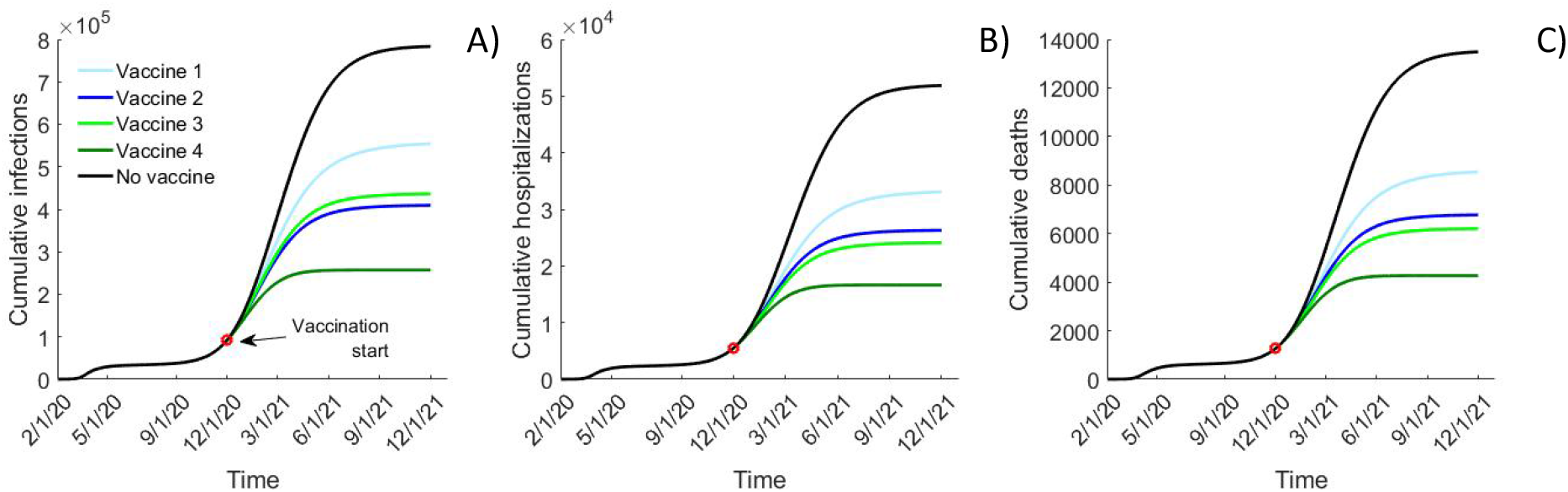
Epidemic projections with different vaccine efficacy profiles. Comparison of A) cumulative infections; B) cumulative hospitalizations and C) cumulative deaths over time simulated with different efficacy profiles assuming that a vaccination start date of Dec. 1, 2020 and rolled out with 5,000 vaccinated daily until 1,000,000 vaccinations are reached. Vaccine 1 (10% VE_SUSC_ and 44.4% VE_SYMP_) and Vaccine 2 (40% VE_SUSC_ and 16.7% VE_SYMP_) result in 50% reduction in symptomatic disease (VE_DIS_) while Vaccine 3 (10% VE_SUSC_ and 88.9% VE_SYMP_) and Vaccine 4 (80% VE_SUSC_ and 50% VE_SYMP_) result in 90% VE_DIS_. All projections represent the mean value of epidemic simulations using 100 calibrated parameter sets.

In comparison, a “susceptibility reducing” vaccine (Vaccine 2: moderate VE_SUSC_, low VE_SYMP_) with the same efficacy against COVID-19 disease (50% VE_DIS_) is expected to prevent an additional 150,000 infections (20% more) and 1800 deaths (15% more) over 1 year (**Fig.2**, Vaccine 2). These differences correspond to 69% and 36% relative increase in the population impact over the projections utilizing the “symptom reducing” (Vaccine 1), respectively.

### Population impact of vaccines reducing the risk of COVID-19 disease by 90%

A “symptom reducing” vaccine with 90% VE_DIS_ (Vaccine 3: low VE_SUSC_, high VE_SYMP_) is expected to have a comparable population impact to the “susceptibility reducing” vaccine with 50% VE_DIS_ (Vaccine 2). Vaccine 3 is projected to have a slightly smaller effect on SARS-CoV2 transmission than Vaccine 2 (51% vs 54% reduction over 1 year) while reducing 2100 more hospitalizations and 550 more deaths than Vaccine 2 (**Fig.2**, light green vs. dark blue).

Finally, a “susceptibility reducing” vaccine with 90% VE_DIS_ (Vaccine 4: high VE_SUSC_, moderate VE_SYMP_) is projected to be the most effective at population level. It is expected to prevent an extra 180,000 infections, 7500 hospitalizations and 1900 deaths over Vaccine 3 over the year after the vaccination start reducing the cumulative infections to approximately 250,000 and cumulative deaths to approximately 4300. (**Fig.2**, Vaccine 4).

**Figure 3** shows the population impacts projected by our model as proportions of infections and deaths prevented, for vaccines with different combinations of effects on susceptibility (VE_SUSC_) and the risk to develop symptomatic disease upon infection (VE_SYMP_) assuming that the vaccine does not reduce the infectiousness for breakthrough infections (VE_INF_=0). The thick lines, displaying the efficacy profiles corresponding to 50% VE_DIS_ and 90% VE_DIS_, demonstrate the wide range of population effectiveness which may result from the same reduction of the risk of COVID-19 disease observed in clinical studies. This uncertainty is particularly large for moderately effective vaccines (50% VE_DIS_) which may result in 30% to 58% reduction in transmission and 38% to 58% reduction in COVID-related mortality depending on which vaccine effects are induced. Vaccines with 90% VE_DIS_ are projected to prevent between 47% and 78% of the infections and between 58% and 77% of deaths over one year. Notably, the effectiveness ranges for moderate and highly effective vaccines overlap which suggest that “susceptibility reducing” vaccine with moderate efficacy against COVID disease may have greater population impact than highly effective “symptom reducing” vaccine. The importance of the effects on the disease severity (VE_SYMP_) decreases if the vaccine provides a strong protection against acquisition (VE_SUSC_) with the contour line corresponding to the same level of impact becoming more horizontal.

**Figure 3.**
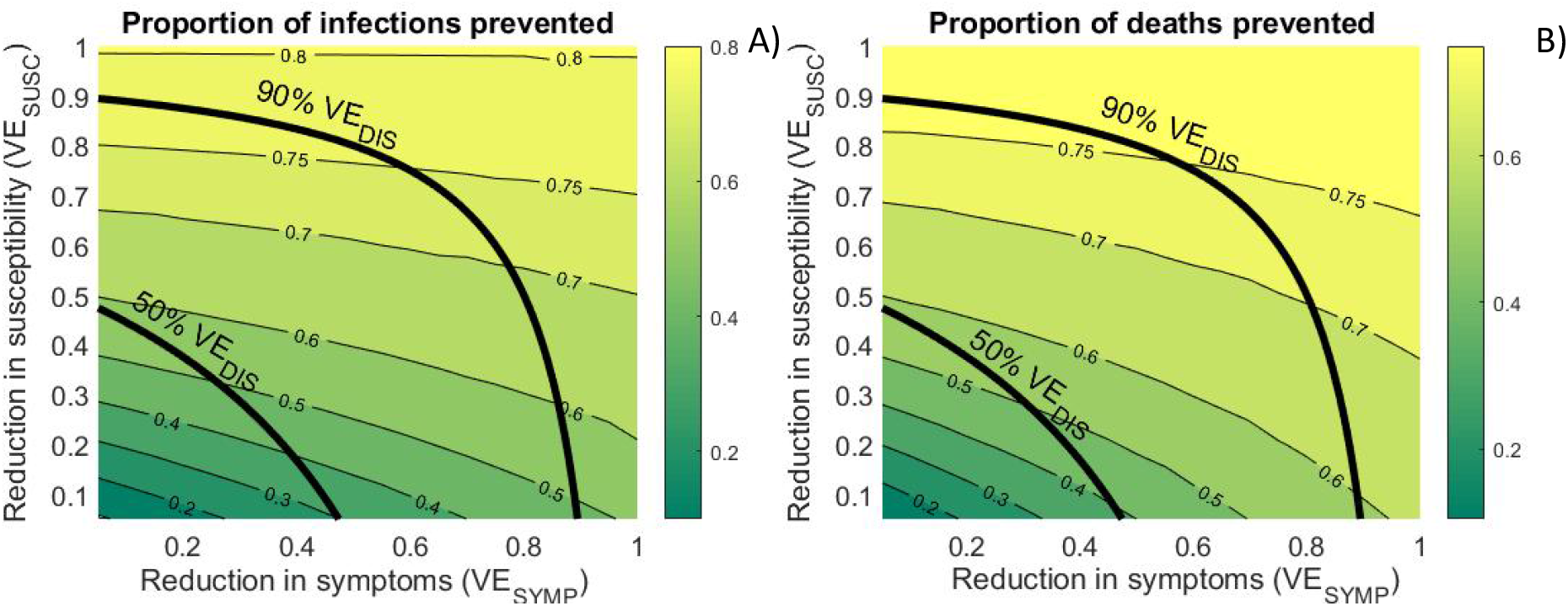
Projected vaccine effectiveness. Contour plots of the proportions of: A) cumulative infections prevented and B) cumulative deaths prevented over 1 year after the start of vaccine rollout by vaccines with different effects on susceptibility (VE_SUSC_) and the risk to develop symptoms (VE_SYMP_). Rollout assumes 5,000 vaccinated daily till 1,000,000 vaccinations are reached. Thick lines represent VE profiles resulting in 50% and 90% reduction in symptomatic disease (VE_DIS_).

### It is critical to rollout the vaccine early and rapidly

We also investigated the importance of the vaccination start date. Our analysis demonstrates that if the vaccine becomes available before the beginning of an outbreak it substantially increases the projected population impact. Based on the simulated epidemics, we show that the lowest effectiveness is recorded in simulations which experience epidemic peak early, within a month after the vaccination start date (**Fig.S5A**, blue). Conversely, simulations with the highest vaccine effectiveness are those in which, in absence of vaccination, cases are expected to peak late, toward the end of projected vaccination rollout (**Fig.S5A**, red). The 25 simulations with the earliest transmission peak (before February 10, 2021) average only 36% and 51% reduction in mortality for the vaccines with 90% VE_DIS_ (Vaccine 3 and Vaccine 4), respectively (**Fig.S5H**,**I**, blue). In comparison, the 25 simulations with latest transmission peak (after April 6, 2021) average 82% and 95% effectiveness in reducing mortality for Vaccine 3 and Vaccine 4, respectively (**Fig.S5H**,**I**, red).

To explore this concept further, we simulated scenarios in which the vaccine start date is delayed by 3 months to Mar 1, 2021. This delay decreases the expected population impact of the vaccination by approximately 40% with all of the simulated VE profiles preventing less than half of the infections, hospitalizations and deaths over 1-year from the start of the rollout (**Fig.4A-C**, gray boxes). Conversely, we estimated that if vaccines had been available even earlier (in Sept 2020) it would have resulted in substantially larger population effectiveness across simulated VE profiles averting 25-30% more infections, hospitalizations and deaths (**Fig.S6**).

We also demonstrate the importance of fast vaccine rollout in response to current concerns that limited quantities of the effective vaccines will be available by late Spring 2021. Simulations with up to 2M vaccinated (more than 90% of the population of King County, WA) project that only 450K vaccinated (∼20% coverage) with the “susceptibility reducing” Vaccine 4 (90% VE_DIS_, high VE_SUSC_) will be enough to reduce the COVID-19 transmission and mortality in half (**Fig 4D-F**, dark green) in conjunction with physical distancing measures to keep physical interactions to 60% of pre-COVID levels (i.e., distancing, masking etc.). In comparison, the use of “symptom reducing” vaccine with the same 90% VE_DIS_ (Vaccine 3) will require 980K vaccinations to reach these thresholds (**Fig 4D-F**, light green). Reducing 80% of the annual transmission is impossible with Vaccine 1-3 even at 90% coverage but is achievable with 1.2M vaccinated (∼55% coverage) with Vaccine 4. Notably, at all coverage levels, the population impact of “susceptibility reducing” Vaccine 2 with moderate 50% VE_DIS_ is comparable with the impact of “symptom reducing” Vaccine 3 with high 90% VE_DIS_.

**Figure 4.**
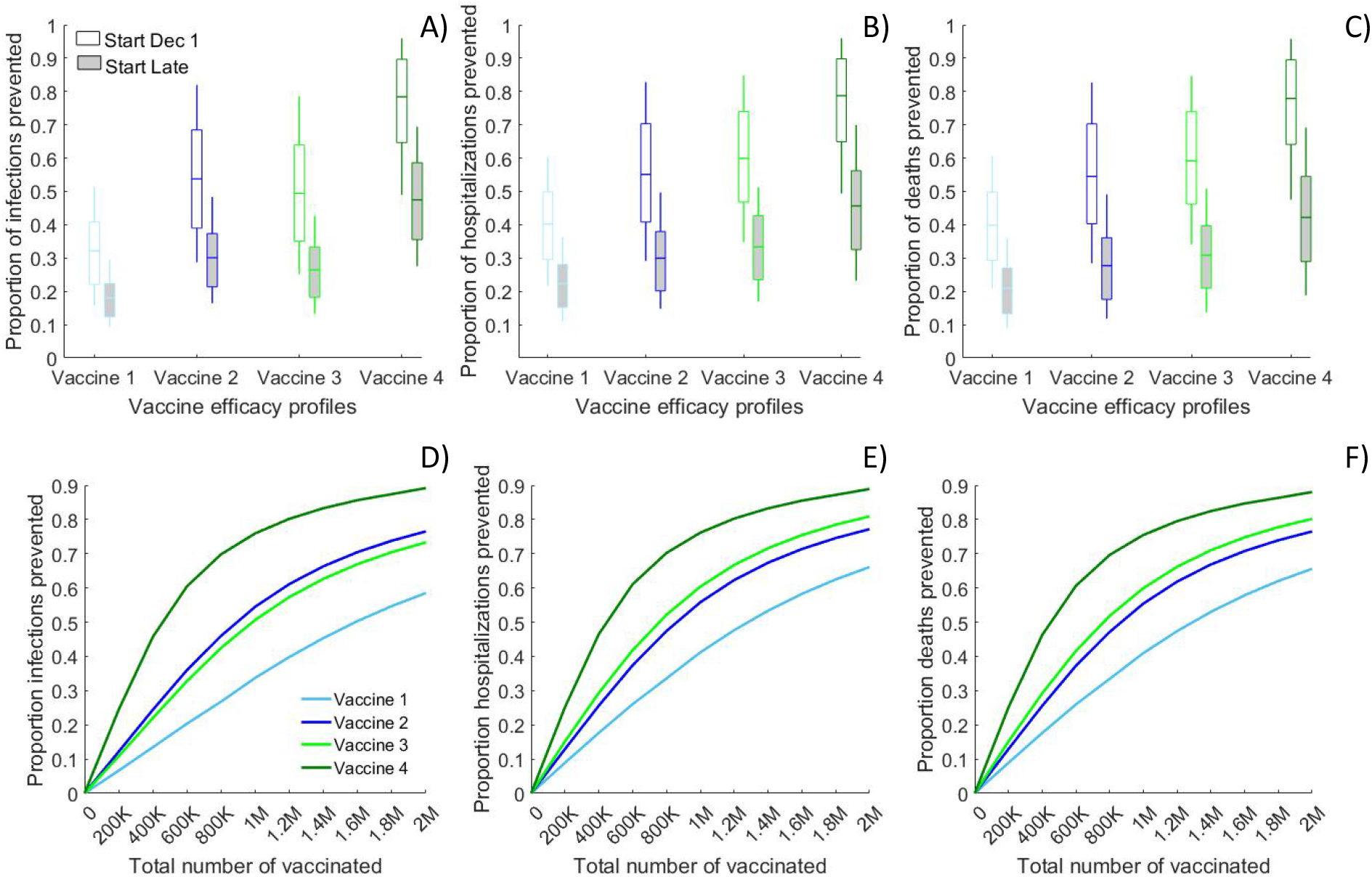
Importance of the timing and the coverage achieved with the vaccine rollout. Comparison of the projected reductions in the number of: A, D) cumulative infections; B, E) cumulative hospitalizations and C, F) cumulative deaths over 1 year due to the use of vaccines with different efficacy profiles. A)-C) compare vaccine rollouts which start on Dec. 1, 2020 (white boxes) or March 1, 2020 (gray boxes) assuming 5,000 vaccinated daily till 1,000,000 vaccinations are reached. D)-F) show the impact of rollouts starting on Dec. 1, 2020 and reaching different number of total vaccinations over 200 days. Vaccine 1 (10% VE_SUSC_ and 44.4% VE_SYMP_) and Vaccine 2 (40% VE_SUSC_ and 16.7% VE_SYMP_) result in 50% reduction in symptomatic disease (VE_DIS_) while Vaccine 3 (10% VE_SUSC_ and 88.9% VE_SYMP_) and Vaccine 4 (80% VE_SUSC_ and 50% VE_SYMP_) result in 90% VE_DIS_. Lines represent the mean value while boxplots represent the uncertainty generated by 100 calibrated simulations.

### When a vaccine rollout may result in small population impact?

The population effectiveness of future vaccine rollouts depends on multiple factors beyond vaccine efficacy profile (VE_SUSC_, VE_SYMP_ and VE_INF_) and the timing of vaccine availability. This includes the speed of the rollout, the vaccination coverage achieved as well as the transmission from contacts with asymptomatic cases. Since our goal is to investigate the minimum impact that can be expected with a licensed vaccine with 50% VE_DIS_ or 90% VE_DIS_ in clinical studies, we simulated a scenario in which: i) no asymptomatic and pre-symptomatic cases are diagnosed, i.e, all SARS-CoV-2 testing is done after symptoms occur and ii) the overall infectiousness of individuals who never express symptoms is only 28% lower than symptomatic cases. Both assumptions reduce the projected impact of all simulated vaccines but especially of Vaccine 1 and Vaccine 3, which mostly shift symptomatic to asymptomatic infections (**Fig. S7**). Under this pessimistic scenario the population effectiveness of “symptom reducing” vaccines is reduced to 8% (Vaccine 1) and 10% (Vaccine 3) when measured as proportion of transmission prevented and to 15%-23% if measured as reduction in mortality. These estimates should serve as a lower bound for the population effectiveness of licensed vaccines with unclear protection against infection. Notably, none of the vaccine profiles projected increased transmission for any of the simulated epidemic conditions even in this pessimistic scenario.

## Discussion

The COVID-19 vaccines currently being tested must lower the number of symptomatic cases by 50% relative to placebo (50% VE_DIS_) in order to be licensed. Recent reports suggest that the first generation of mRNA vaccine have remarkable VE_DIS_ but their ability to prevent infection is currently not a criterion for approval and is unknown. Ultimately, seroconversion data may provide some insight regarding breakthrough asymptomatic infection but may be marked by loss of a humoral response such that underestimation of asymptomatic infection is possible.

In this analysis we explored different combinations of vaccine effects which result in 50% and 90% reduction of symptomatic COVID-19 and demonstrated significant variability in projected population effectiveness based on whether the vaccines fully prevent infection or convert symptomatic cases to asymptomatic cases with some ongoing possibility of transmission. This result highlights the potent indirect effects that may accrue from stopping further chains of transmission and underscores the need to discriminate VE_SUSC_ from VE_SYMP_. Although this difference is not being evaluated as a primary endpoint in current studies, estimating the frequency and downstream transmission likelihood of asymptomatic infection is recognized as a research priority.^12,38,39^ We at least identify that a high rate of conversion to asymptomatic infection is unlikely to increase the incidence of new cases. Vaccines with high VE_SUSC_ are expected to provide larger population benefits in terms of preventing hospitalizations and deaths. We demonstrate that the population effectiveness of “susceptibility reducing” vaccines can be up to 70% larger than “symptom reducing” vaccines even if resulting in the same observed VE_DIS_. Such vaccines contribute to a more rapid attainment of necessary vaccination thresholds to achieve herd immunity. Moreover, we found that a “susceptibility reducing” vaccine with moderate 50% VE_DIS_ may provide comparable the population benefits with a “symptom reducing” vaccine with high 90% VE_DIS_. With multiple vaccine candidates in the pipeline, our results reveal the complexity of the problem and suggest that the usefulness of a vaccine should not be judged by a single efficacy number.

A significant proportion of the population impact expected with a “symptom reducing” vaccine can be attributed to the reduced infectiousness of the asymptomatic cases as has been reported for influenza vaccine.^40^ This is despite the fact that asymptomatically infected people may remain infectious for a longer period of time given the lower likelihood of being diagnosed and self-isolating. Our analysis suggests that the population effectiveness of “symptom reducing” vaccine, measured as reduction in SARS-CoV2 transmission, may decrease to below 10% if the infectiousness gap between asymptomatic and symptomatic cases is smaller and if individuals who don’t show symptoms are not readily tested, diagnosed and isolated. This highlights the need to continue the contact tracing and quarantine measures, currently in place, for at least the duration of the vaccine rollout.

Another consideration is that vaccine effects on disease progression, including reduced severity and symptom manifestation, could be correlated with vaccine effects on the infectiousness of the breakthrough infections if both effects result from reduction in the peak viral load. If this is the case, the projected impact of “symptom reducing” vaccines here could be underestimated. None of the ongoing clinical studies are designed to answer these questions. The ideal studies to address measurement of VE_INF_ would measure secondary attack rates of household members of vaccine versus placebo recipients or would measure viral loads carefully among infected vaccine and placebo recipients if exposure viral load is identified as a validate surrogate of transmission likelihood.

We also conclude that vaccine rollout will be significantly more effective if introduced prior to rather than during a surge in cases. Even the rollout of a vaccine with 90% VE_DIS_ may result in very small reduction in actual SARS-CoV-2 transmission if it occurs after a substantial number of people have been already infected. Unfortunately, at the time of this writing, the effective reproductive number exceeds one in virtually every U.S. state and many hospitals are already nearing capacity in many regions of the country. During the current surge, there is little evidence as of yet that case incidence has started to lower as states in the Northern Plains now have an estimated prevalence approaching 25% and few US states have reimposed lockdowns.^41,42^ On November 15, 2020 Washington state (and thus King County, the focus of this paper) has reimposed some restrictions closing businesses and limiting gatherings but the impact of these measures has not affected the upward epidemic trajectories yet.^43^ If the current uncontrolled increase in confirmed cases and hospitalizations across the U.S. continues, the expected vaccination in Spring 2021 may come too late. The most recent projections of the Institute for Health Metrics and Evaluations predict that the trend of increasing cases in Washington State will continue until February 2021^42^. Our analysis provides a strong rationale for mitigation measures such as mask mandates^44^ and further physical distancing^45,46^ to contain the epidemic surge and weather the incoming months in expectation of mass production and distribution of safe and effective vaccines. The importance of an early vaccine rollout has also been highlighted in other studies.^20^

Our results qualitatively agree with previous studies^18,24^ that supporting non-pharmaceutical measures might be needed during vaccination campaigns to slow down the epidemic and increase its effectiveness. We also share the conclusion of other modeling teams that vaccines mediated by a reduction in susceptibility to infection will have a greater impact than disease-modifying vaccines.^25,26^ However, our analysis suggests that mortality reduction greater than 80%, projected in some studies^24^, is achievable under moderate vaccine coverage (40%-50%) only if a “susceptibility reducing” vaccine is rolled out rapidly in well-controlled epidemic environment but not in the middle of an epidemic wave.

Some caveats of this work are important to note. First, we assumed that the licensed vaccines have no direct effect on the infectiousness (VE_INF_) of the breakthrough infections. Nevertheless, the vaccine shifting symptomatic infections to less infectious asymptomatic infection indirectly reduces overall infectiousness. Adding a non-zero VE_INF_ to the vaccine efficacy profile would improve the estimated population impact, so our projections are therefore conservative and present a worst-case scenario. While a moderate VE_INF_ will not directly protect the vaccine recipient, it would likely effectively limit further chains of transmission and therefore should be evaluated. Second, we don’t explicitly model two-dose vaccine regimens but assume immediate immunization at the time of vaccination. Therefore, the vaccine effects before the full regimen is given, i.e. before the second dose for 2-dose regimens, are ignored. Third, we considered vaccination rollouts where all age groups are vaccinated. Current recommendations point to vaccinating older age groups first, which are those at higher risk. In that sense, our model might be underestimating the impact of vaccination rollouts on the number of deaths prevented. Finally, we assume that the level of physical interactions remains steady for the next year which results in epidemic outbreaks of various magnitudes providing a reasonable variability in the counterfactual epidemic conditions in which we evaluate vaccines with different efficacy profiles. Enforcing partial or full lockdowns and reopenings would likely result in periodic fluctuations in the number of infections and confirmed cases^47^ but unlikely to affect effectiveness projections if the same schedule is applied to scenarios with and without vaccination. However, if physical interactions increase with the rollout of vaccines either due to policy shifts or individual behavioral compensation, the resulting outcomes would be worse than what we have predicted here.

The results of this analysis suggest that fully understanding the profile of the vaccine is important, since vaccines with different profiles may show similar efficacy in clinical studies but have considerably different population impact. In particular, vaccines which prevent COVID-19 disease but not SARS-CoV-2 infection, and thereby shift symptomatic infections to asymptomatic infections, will prevent fewer infections and require larger and faster vaccination rollouts to achieve the same population impact as vaccines that reduce susceptibility to infection.

## Supporting information

Technical Supplement

## Data Availability

All data and code used for analysis are available upon request from the corresponding author

