## Supplementary material for "COVID-19 vaccines that reduce symptoms but do not block infection need higher coverage and faster rollout to achieve population impact": Technical Supplement

### 1. Complete model description

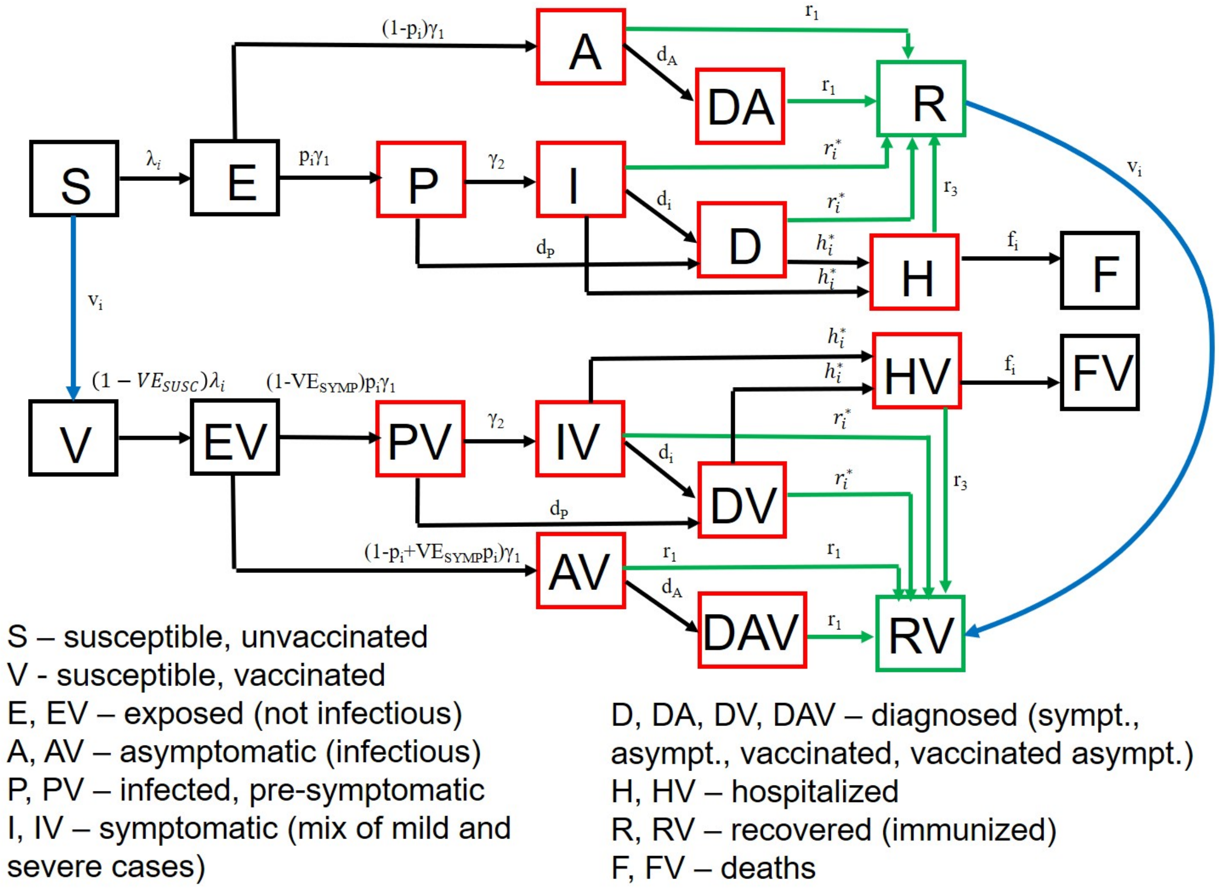


**Figure S1. Model diagram** Structure of the mathematical model of SARS-CoV-2 dynamics. Stratification by age (4 groups) is not shown. Red compartments are infectious while green compartments are recovered with long-lasting natural immunity. Blue arrows indicate vaccination.

**The model is described by a set of differential equations for each age group (i=1 for age 0-19 years, i=2 for age 20-49 years, i=3 for age 50-69 years, and i=4 for age 70+ years):**

$$\frac{dS_i}{dt} = -\lambda_i S_i - \frac{v_i S_i}{S_i + R_i}$$

$$\frac{dE_i}{dt} = \lambda_i S_i - \gamma_1 E_i$$

$$\frac{dA_i}{dt} = (1 - p_i)\gamma_1 E_i - (r_1 + d_A)A_i$$

$$\frac{dDA_i}{dt} = d_A A_i - r_1 DA_i$$

$$\frac{dP_i}{dt} = p_i \gamma_1 E_i - (\gamma_2 + d_P)P_i$$

$$\frac{dI_i}{dt} = \gamma_2 P_i - d_i(t) I_i - h_i^* I_i - r_i^* I_i$$

$$\frac{dD_i}{dt} = d_P P_i + d_i(t) I_i - h_i^* D_i - r_i^* D_i$$

$$\frac{dH_i}{dt} = h_i^* I_i + h_i^* D_i - r_3 H_i - f_i \quad H_i$$

$$\frac{dF_i}{dt} = f_i \quad H_i$$

$$\frac{dR_i}{dt} = r_1 A_i + r_1 DA_i + r_i^* \quad I_i + r_i^* D_i + r_3 H_i - \frac{v_i R_i}{S_i + R_i}$$

$$\frac{dV_i}{dt} = -(1 - VE_{SUSC})\lambda_i V_i + \frac{v_i S_i}{S_i + R_i}$$

$$\frac{dEV_i}{dt} = (1 - VE_{SUSC})\lambda_i V_i - \gamma_1 EV_i$$

$$\frac{dAV_i}{dt} = (1 - p_i + VE_{SYMP} p_i)\gamma_1 EV_i - (r_1 + d_A)AV_i$$

$$\frac{dDAV_i}{dt} = d_A AV_i - r_1 DAV_i$$

$$\frac{dPV_i}{dt} = (1 - VE_{SYMP}) p_i \gamma_1 EV_i - (\gamma_2 + d_P)PV_i$$

$$\frac{dIV_i}{dt} = \gamma_2 PV_i - d_i(t) IV_i - h_i^* IV_i - r_i^* IV_i$$

$$\frac{dDV_i}{dt} = d_P PV_i + d_i(t) IV_i - h_i^* DV_i - r_i^* DV_i$$

$$\frac{dHV_i}{dt} = h_i^* IV_i + h_i^* DV_i - r_3 HV_i - f_i \quad HV_i$$

$$\frac{dFV_i}{dt} = f_i \quad HV_i$$

$$\frac{dRV_i}{dt} = r_1 AV_i + r_1 DAV_i + r_i^* \quad IV_i + r_i^* DV_i + r_3 HV_i + \frac{v_i R_i}{S_i + R_i}$$

$p_i$ – proportion of the infections which become symptomatic by age in absence of a vaccine

$\gamma_1, \gamma_2$- progression rates from exposed (E) to infectious (A and P) to symptomatic (I)

$h_i$ – hospitalization rate among severe cases by age in absence of a vaccine

$h_i^*$ – hospitalization rate among diagnosed by age (calculated)

$r_{1\text{-}3}$ - recovery rate of the asymptomatic, mild symptomatic and hospitalized cases

$r_i^*$ - recovery rate of the diagnosed symptomatic cases by age (calculated)

$f_i$ – fatality rate among hospitalized by age

$VE_{SYMP}$ – vaccine efficacy in reducing the risk of symptomatic disease after infection

$d_i$ – diagnostic rate by age. They vary in time being initially set at zero and later elevated after the start of the COVID measures at t=$\delta_1$. Finally, they are further elevated after testing capacity was increased t=$\delta_3$, i.e. $d_i(t) = \begin{cases} 0, & t < \delta_1 \\ d_{i,1}, & \delta_1 \le t < \delta_3 \\ d_{i,2}, & t \ge \delta_3 \end{cases}$

The forces of infection ($\lambda_i$), representing the risk of the susceptible individuals by age to acquire infection (transition from susceptible to exposed), are differentiated by age of the susceptible individual, the contact matrix (proportion of contacts with each age group), infection and treatment status (asymptomatic, pre-symptomatic, symptomatic, diagnosed and hospitalized cases) of the infected contacts, and the time-dependent reduction of transmission due to physical distancing measures (work from home, closing non-essential businesses, banning large gathering, etc,) applied in the area (scaled up starting March 8 and fully taking effect March 29) and later relaxed during the reopening after May 15.

$$\lambda_i = \sum_{j=1}^{4} c_{ij}\left(1 - R_{sd}(t)\right)\big[\beta_a A_j + \beta_p P_j + \beta_s I_j + \beta_d D_j + \beta_{da} DA_j + (1 - VE_{INF})\left(\beta_a AV_j + \beta_p PV_j + \beta_s IV_j + \beta_d DV_j + \beta_{da} DAV_j\right)\big]/N_j + c_{ij}\beta_h(H_j + HV_j)/N_j$$

$$\lambda_{vi} = (1 - VE_{SUSC})\lambda_i$$

where

$\beta_a$, $\beta_p$, $\beta_s$, $\beta_d$, $\beta_h$ are the transmission rates from contacts with asymptomatic, pre-symptomatic, symptomatic, diagnosed and hospitalized infections (before the start of COVID measures at t= $\delta_1$),

$c_{ij}$ – contact matrix (proportion of the contact with other age groups),

$N_i$ – population size by age,

$VE_{SUSC}$ – vaccine efficacy in reducing susceptibility to infection

$VE_{INF}$ – vaccine efficacy in reducing the infectiousness

$R_{sd}$ (t) is the reduction of transmission due to physical distancing and other preventive measures which is applied uniformly to all age groups. It is scaled up linearly from 0 to $R_{sd}^{max}$ between t= $\delta_1$ and t= $\delta_2$). Later it is decreased for all age groups (in the baseline scenario) or only age groups 1-3 (in the Protect Seniors scenario) from $R_{sd}^{max}$ to some pre-defined value during reopening period (between t= $\delta_3$ and t= $\delta_4$), It is further reduced to 20% for the youngest group at school reopening (t= $\delta_5$).

### 2. Model parameterization

**Table S1. Parameters and ranges used in the analysis. (Fixed in black, Scenarios in blue, Calibration in red)**

| Parameter | Description | Values and ranges | Type |
|---|---|---|---|
| $\gamma_1$ | **Progression rates from exposed (E) to infectious (A or P) (latent time)$^{-1}$** | **(3 days)$^{-1}$** | Fixed |
| $\gamma_2$ | **Progression rates from pre-symptomatic (P) to symptomatic (I) (pre-symptomatic time)$^{-1}$** | **(2 days)$^{-1}$** | Fixed |
| $p_i$ | **Proportion of the infections which become symptomatic by age** | **80%** | **Fixed** |
| $v_i$ | **Vaccination rate by age. Number of currently uninfected individuals who get vaccinated daily for 200 days after the start of vaccination** | Proportional by age-group size (see Table S3)<br>Main: 5000<br>Alternative: 1000-10000 | **Scenarios** |
| $d_{i,j}$ | **Diagnostic rate if symptomatic by age (*i*) after COVID measures are initiated (*j=1*) and after testing capacity was increased (*j=2*)** | **$d_{i,1}$=1-10%**<br>**$d_{i,2}$=10%** | Calibrated |
| $d_A, d_P$ | **Diagnostic rate if asymptomatic and pre-symptomatic before (*j=1*) and after (*j=2*) testing capacity was increased** | **$d_{A,1}$= $d_{P,1}$=0**<br>***Main:* $d_{A,2}$= $d_{P,2}$=5%**<br>***Alternative:* $d_{A,2}$= $d_{P,2}$=0** | Scenarios |

| | | | |
|---|---|---|---|
| ***id*** | **Symptomatic infectiousness duration** | **7 days** | Fixed |
| $\beta_i$ | **Daily transmission from infected from asymptomatic, pre-symptomatic, symptomatic, diagnosed and hospitalized groups in absence of COVID measures** | ***=βs*(1, $\beta_p$, 1, $\beta_d$, 0)***<br>$\beta_p$ calculated to get 44% pre-sympt. transmission<br>***$\beta_s$ =R0/(βp/$\gamma_2$ + id) for R0 = 2.2-4***<br>***βd =0.5-0.75 (Lockdown)***<br>***Decreased 50% afterward*** | Calculated |
| $R_{sd}^{max}$ | **Maximal reduction of transmission due to social distancing (scaled up linearly between t= $\delta_1$ and t= $\delta_2$)** | ***50%-90%*** | Calibrated |
| $\delta_0$ | **Number of days between the start of the simulation (day 0) and the 1st diagnosed case from data (Feb.28)** | ***40-50*** | Calibrated |
| **[δ1, δ2]** | **Period of scaling up COVID measures** | **March 8-29** | Fixed |
| **[$\delta_3$, $\delta_4$]** | **Reopening period** | **May 15- July 15** | Fixed |
| $m_i$ | **Proportion of symptomatic case which remain mild by age** | **99.8%, 96.9%, 87.1%, 74.5%** | Fixed |
| $h_i$ | **Hospitalization rate among severe cases by age** | ***$h_1$=0.1, $h_2$=0.15, $h_3$=0.15-0.3, $h_4$=0.15-0.3*** | Calibrated |
| $h_i^*$ | **Hospitalization rate among symptomatic and diagnosed cases** | $= (1 - m_i)h_i$ | Calculated |
| $r_1$ | **Recovery rate of asymptomatic cases** | ***Main: 1/id,***<br>***Alternative: 1/9*** | Scenarios |
| $r_2$ | **Recovery rate of mild symptomatic cases** | ***1/id*** | Fixed |

| | | | |
|---|---|---|---|
| $r_i^*$ | **Recovery rate of the symptomatic and diagnosed cases** | $= m_i r_2$ | Calculated |
| $r_3$ | **Recovery rate of the hospitalized cases** | ***1/14*** | Fixed |
| ***hd*** | **Time from hospitalization to death** | **11.2 days** | fixed |
| $f_i$ | **Fatality rate by age among hospitalized before reaching ICU capacity* (overall mortality when hospitalized/time to death) adjusted for underreporting of mild cases** | **=α*CFR/(1-m$_i$)/hd**<br>**CFR: (0%,0.2%,2.1%,15.9%)**<br>**α =0.8-1.25** | Calibrated |

**Table S2. Contact matrix**. The columns represent the distribution of contacts of a person from given age group across all age groups:

| Proportion contacts with | 0-19 y | 20-49 y | 50-69 y | 70+ y |
|---|---|---|---|---|
| 0-19 y | 0.56 | 0.24 | 0.15 | 0.18 |
| 20-49 y | 0.34 | 0.57 | 0.49 | 0.34 |
| 50-69 y | 0.08 | 0.16 | 0.29 | 0.28 |
| 70+ y | 0.01 | 0.03 | 0.07 | 0.20 |

**Table S3. King County age pyramid based on data from 2017**

| Proportion of the population | 0-19 y | 20-49 y | 50-69 y | 70+ y |
|---|---|---|---|---|
| | 22.93% | 45.52% | 23.50% | 8.05% |

#### 3. Model Calibration

The model is calibrated to 5 “targets” based on local data **(Fig 2)**, including: i) overall number of confirmed cases (target #1) and deaths (target #2) reported in King County over time since the start of the epidemic outbreak through April 30; ii) age-distribution of the cumulative confirmed cases (target #3) and deaths (target #4) reported in King County at 3 time points after the start of the epidemic outbreak and iii) the timing of the peak of daily confirmed cases (target #5) estimated as April 1. We used a genetic algorithm (NSGA-II multivariate optimization algorithm in the mco R package) to evolve a population of parameterizations to arrive at a set approximating the Pareto front. We defined thresholds for each target, based on a reasonable fit to that target and used them to select the best fit from the final population.
Next, we used Monte Carlo filtering to select 100 parameter sets which reproduce data within these pre-specified tolerances. Resulting sets were used to explore the uncertainty in our model projections (see **Fig S2**).

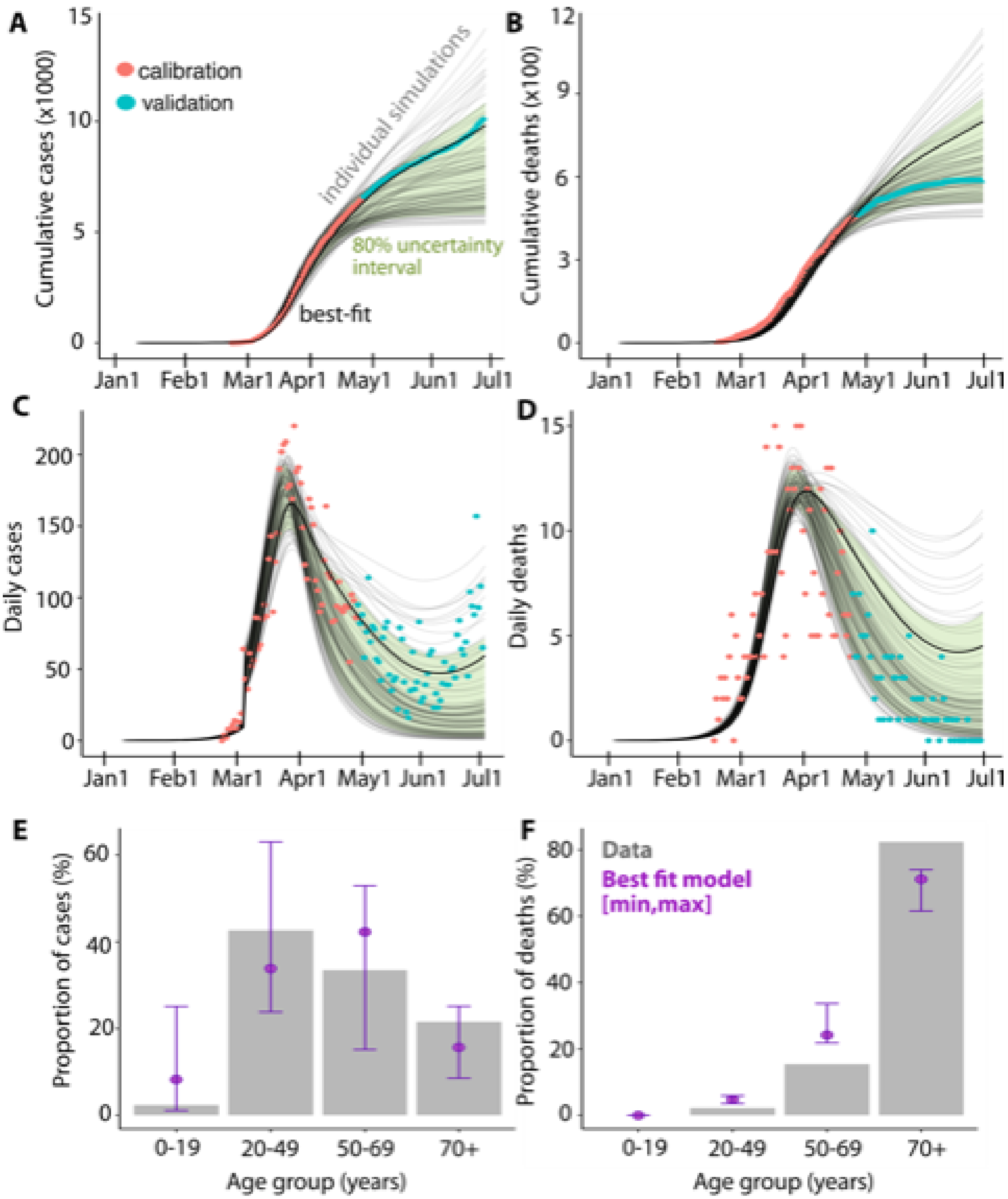


**Figure S2. Model calibration and validation.** Model fitting to 5 sources of King County data assuming gradual scale up of social distancing between March 8 and March 29: A)-B) Cumulative and daily cases and deaths. Red dots represent data up to April 30, thick lines represent the best model fit while other acceptable trajectories are shown in grey. Green bands show 80% range from acceptable trajectories. C)-D) Age distributions of cases and deaths as of April 15. Bars represent data, green dots and ranges represent the best fit and other acceptable trajectories included in the analysis. Reopening plan is implemented between May 15 and July 15 by gradually restoring 60% of pC_PI in all age groups.

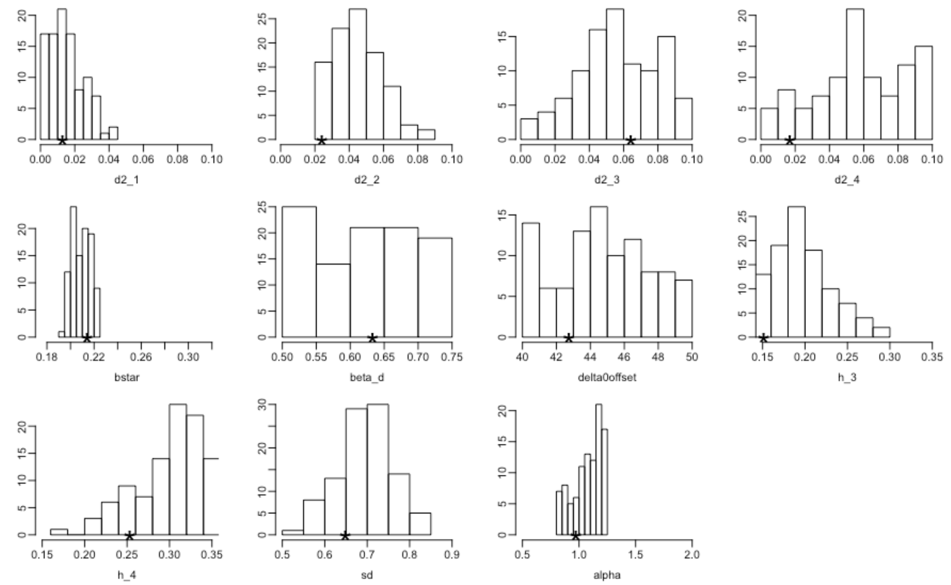


**Figure S3. Calibrated parameter sets**. Shown are the distributions of the parameter values for the 100 acceptable parameterizations (UI) with a * marking the value of the 'best' parameterization.

We validate our population model by predicting independent data not used for calibration: i) cumulative number of confirmed cases and deaths between April 30 and June 30, as well as estimated number of daily hospitalizations and the overall hospitalization rates among confirmed cases at the end of April; ii) expert predictions informed by seroprevalence data of the cumulative incidence. We also compare to independent region-specific modeling projections including cumulative SARS-CoV-2 incidence at the beginning of March based on genomic analyses[1] as well as the cumulative incidence and effective reproductive number (Rt) estimates for King County for the period between March 1 and April 22 reported by the Institute for Disease Modeling[2].

### 4. Additional results

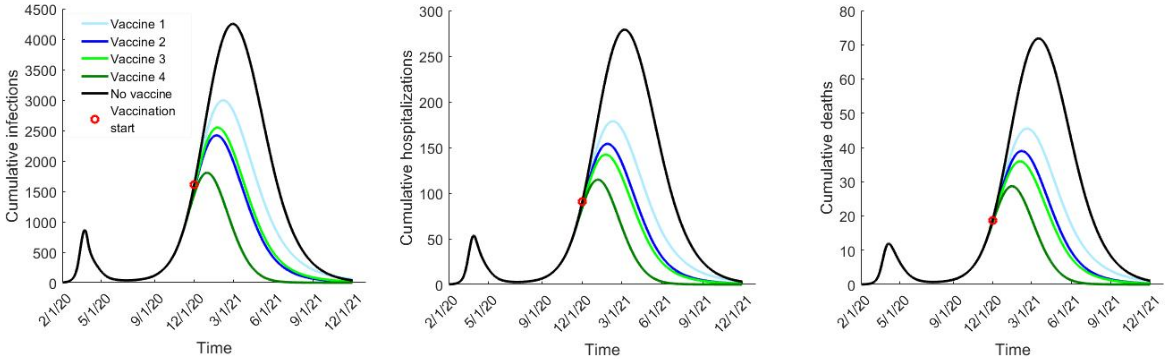

**Figure S4. Average daily transmission and mortality curves based on simulations with calibrated parameter sets.** Vaccine 1 and 2 result in 50% reduction in symptomatic disease ($VE_{DIS}$) while Vaccine 3 and 4 result in 90% $VE_{DIS}$. Vaccine rollout begins on Dec.1 with 5,000 vaccinated daily till 1,000,000 vaccinations are reached

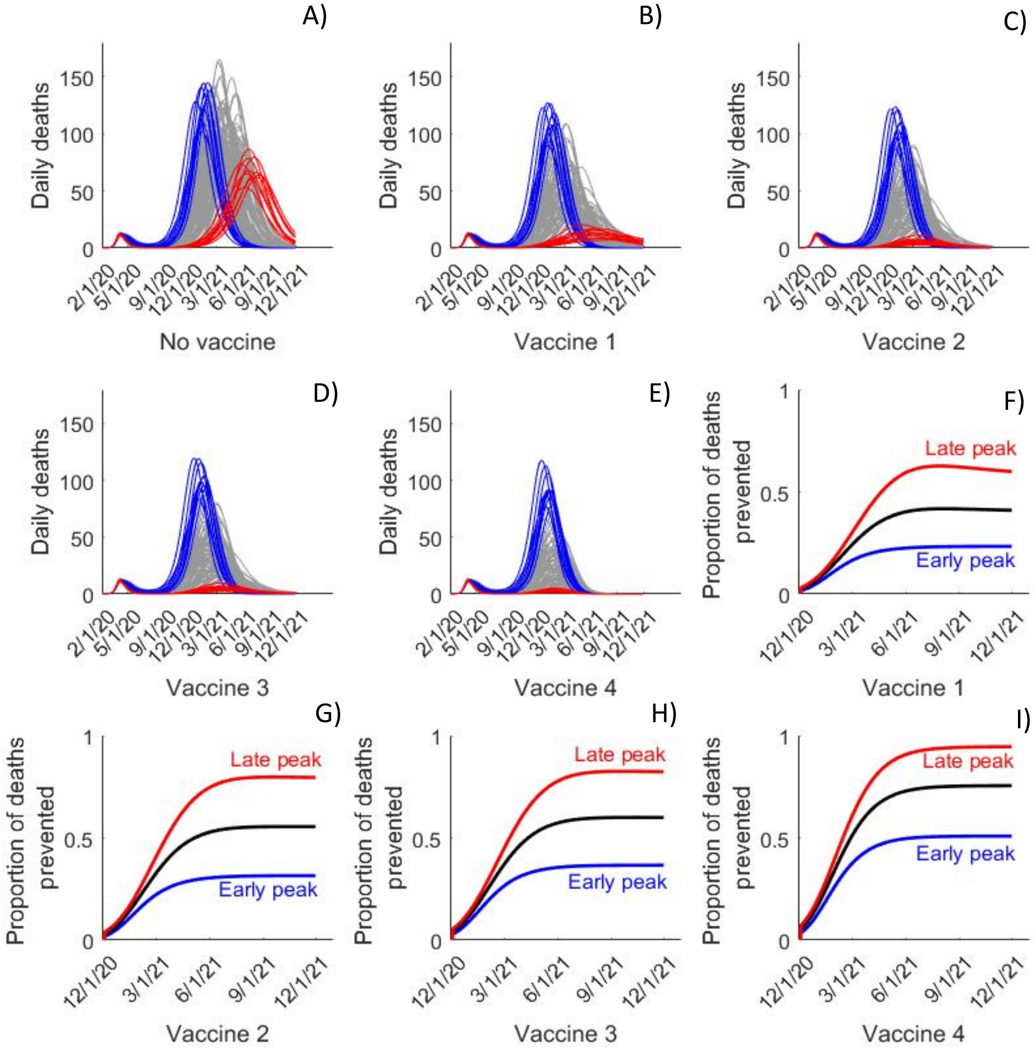


**Figure S5 Association between outbreak transmission peak and vaccine effectiveness** A)-E**)** Daily transmission and mortality curves for the 100 selected simulations. Simulations with the 10 highest reductions of mortality are colored in red while the 10 lowest are colored in blue. F)-I) Projected vaccine effectiveness based on all simulations (black), only the 25 simulations with the earliest transmission peak (blue) or the 25 simulations with latest transmission peak (red). Vaccine 1 and 2 result in 50% reduction in symptomatic disease ($VE_{DIS}$) while Vaccine 3 and 4 result in 90% $VE_{DIS}$. Vaccine rollout begins on Dec.1 with 5,000 vaccinated daily till 1,000,000 vaccinations are reached

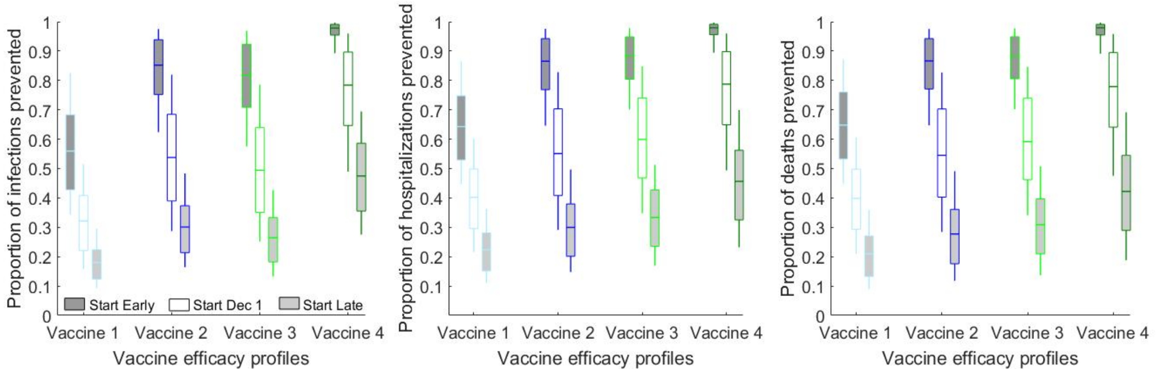


**<u>Figure S6.</u> Importance of the early start of the vaccination campaign.** Comparison of the projected reductions in the number of: A) cumulative infections; B) cumulative hospitalizations and C) cumulative deaths due to the use of vaccines with different efficacy profiles assuming that the vaccines were available on Dec. 1 (white boxes), three months earlier (dark gray boxes) or three months later (light gray boxes). Vaccine rollout assumes 5,000 vaccinated daily till 1,000,000 vaccinations are reached. Vaccine 1 and 2 result in 50% reduction in symptomatic disease ($VE_{DIS}$) while Vaccine 3 and 4 result in 90% $VE_{DIS}$. Boxplots represent the uncertainty generated by 100 epidemic simulations selected at model calibration.

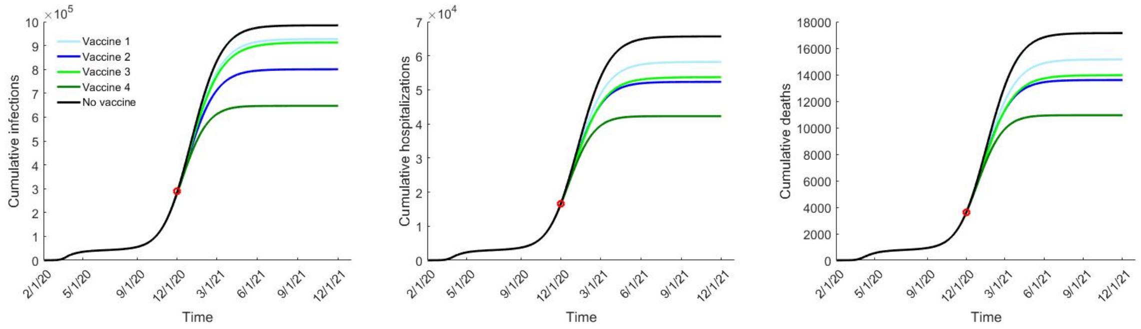


**<u>Figure S7.</u> What is the minimum impact to be expected with a licensed vaccine?** Comparison of: A) cumulative infections; B) cumulative hospitalizations and C) cumulative deaths over time simulated with different efficacy profiles assuming that <u>no asymptomatic and pre-symptomatic cases are diagnosed and the overall infectiousness of individuals who never express symptoms is only 28% lower than symptomatic cases</u>. In all scenarios, the vaccination starts on Dec. 1 and rolled out with 5,000 vaccinated daily till 1,000,000 vaccinations are reached. Vaccine 1 and 2 result in 50% reduction in symptomatic disease ($VE_{DIS}$) while Vaccine 3 and 4 result in 90% $VE_{DIS}$. All projections represent the mean value from 100 epidemic simulations selected at model calibration.
